# “Side effects of Vero cell vaccination against Covid-19 among medical students of Nepalgunj Medical College”-A Post Vaccination survey

**DOI:** 10.1101/2022.12.22.22283763

**Authors:** KC Rupak, Sibika Malla, Aashish Pandey, Merina Shrestha, Saharoj Siddiqui, Suraj Adhikari, KC Niranjan, KC Rumi

## Abstract

**Background and aims:** Severe Acute Respiratory Syndrome Corona Virus 2 (SARS-CoV-2) is a highly transmissible virus causing Coronavirus disease (COVID-19). Its symptoms include fever, dry cough, and shortness of breath. Vaccine plays a significant role in controlling infectious disease and lesser health resources are used, and there is increased allocation of those resources for disease management. Vero Cell is an inactivated vaccine against COVID-19 manufactured by Sinopharm Company of China and recommended for people above 18 years by the World Health Organization. It is administered in two doses of 0.5 ml, 14-28 days apart. The goal of our study was to determine post-vaccination side effects of both the first(1^st^) and the second(2^nd^) dose among participants of Nepalgunj Medical College Teaching Hospital (NGMCTH), Kohalpur, Banke, Nepal.

**Methods:** A prospective cross-sectional study was conducted among undergraduate medical students and intern doctors of Nepalgunj Medical College. A comprehensive structured questionnaire was designed and distributed following each dose of vaccination. Data were collected from 6^th^ January 2022 to 6^th^ April 2022 and entered in Microsoft Excel and analyzed using the statistical package for social sciences (SPSS) version 22. Analyzed data were presented using simple descriptive statistics with appropriate tables.

**Results:** Out of 156 participants, the majority were males (66.7%), within the age group of 21-24. The most frequently encountered symptom was pain at the injection site, predominantly seen in males. Among all the tested variables, the second dose of vaccination showed a significant association with sex. All the predominant symptoms exhibited a significant association following vaccination, except for pain at the injection site (p-value <0.05).

**Conclusions:** Vaccine has proven to be a lifesaving breakthrough during the peak of the COVID-19 outbreak and has been precisely efficacious in a developing country like Nepal.

## INTRODUCTION

The virus that is the root cause of the highly threatening global pandemic has been designated as severe acute respiratory syndrome coronavirus 2 (SARS-CoV-2) causing the coronavirus disease (COVID-19). (1)As per the reports, this virus was first discovered in persons exposed to a seafood wholesale wet market in Wuhan, China. (2)By the 31st of December, the Wuhan Municipal Health Commission had given out its alert. (3)Soon this disease enveloped the entire world and was announced as a pandemic by the World Health Organization on the 11th of March, 2020. (4)Till February 3, 2022, a total of 385,967,643 cases of COVID-19 have been confirmed out of which 5,721,171 has died and 305,840,220 has been recovered. (5)

SARS-CoV-2 is the third coronavirus after SARS-CoV (2002) and MERS-CoV (2012) which has a high case fatality rate of 4.8%. (6,7)It is a single-stranded, spherical, or pleomorphic enveloped RNA virus whose club-shaped protrusions cover them like a crown and this is also the reason for its naming. (8,9) SARS-CoV-2 can be transmitted through direct contact, air, fomite, or fecal-oral route. Similarly, blood-borne, mother-to-child, or animal-to-human transmission of this virus has also been found. (10) Its key symptoms include fever, dry cough, and shortness of breath. Pneumonia is a major complication. (11)

The first positive case of COVID-19 in Nepal was confirmed in a girl who returned from Wuhan Institute of Technology, Wuhan, China on the 13th of January, 2020. (12) As of 12 February 2021, the total confirmed cases there were 972,632 out of which 11,882 had died. (5) The only hope in this dark phase is the vaccine that can end the pandemic. Safe and effective vaccines are a game-changing tool. Vaccine plays a significant role in controlling infectious disease, reduction of the use of health services because of infectious diseases, and leaving more resources for the prevention and treatment of other diseases. (13) It prevents people from being infected or getting gravely sick. It prevents the person from spreading COVID-19 to others, thus making it difficult for the disease to escalate, contributing to herd immunity. (14,15) Till 2 February 2022, 33 vaccines have been approved for clinical use and 65 vaccines are in their phase three trials. (16) The vaccination program has begun in Nepal on 27th January 2021 after India provided one million doses of the Covid-19 Covishield vaccine, developed by the University of Oxford and pharmaceutical giant AstraZeneca and locally produced by the Serum Institute of India, the world’s largest vaccine company. (17) At the beginning of March, COVAX, an international vaccine-sharing scheme supported by the United Nations, supplied 348,000 doses of Covishield. Beijing delivered 800,000 doses of Vero Cell under grant assistance at the end of March. Again, in June, an additional 1 million doses of Vero Cell under a Chinese grant then arrived Nepal. (18)

The Vero cell is an inactivated vaccine manufactured by Sinopharm Company of China which is recommended safe to be used for people above 18 years by the World Health Organization. It is administered in two doses of 0.5 ml each. The second dose is administered 14-28 days after the first one. (19) According to the global health agency’s estimation, the Vero cell’s overall efficacy is about 78%. (20) Sinopharm (Vero Cell) vaccine has been evidently proven to be 50.4% effective in preventing Covid-19 and 94% effective in preventing deaths. (21)

Many people who have been vaccinated are experiencing side effects in one form or the other. Outside of clinical trials situation, only a few data are available which has assessed its safety. This research aims to assess the post-vaccination side effects of Vero Cell (Sinopharm) in undergraduate medical students and medical interns of Nepalgunj medical college.

## METHODS

### Study design and settings

A prospective cross-sectional study was conducted among undergraduate medical students and intern doctors of Nepalgunj Medical College after receiving ethical approval from the institutional review committee of the college. The confidentiality of all the participants was taken into account. Participants promptly received the 1^st^ dose and when the second dose became accessible, our vaccination was completed upon receiving the 2^nd^ dose. The study period spanned from 6^th^ January 2022 to 6^th^ April 2022, a period of 3 months.

### Study Participants

156 participants were included in the study constituting M.B.B.S students from 3^rd^ year to final year and Intern Drs of Nepalgunj medical college, Banke, Nepal.

### Eligibility criteria

Participants who received two doses of the vaccination were eligible to participate in the study. Those participants who didn’t provide their consent for the study and those who have previously been vaccinated with other vaccines like Covishield, Pfizer; etc., and students from basic science were excluded.

### Questionnaire administration

A comprehensive structured questionnaire was designed and distributed among the participants following each dose of vaccination, capturing the demographic information and the post-vaccination symptoms. Subsequently, the results were collected and analyzed two weeks later. Recall bias and information bias may include.

### Statistical Analysis

Data were collected using a comprehensive structured questionnaire. The eligible participants were given the questionnaire twice, after administering each dose of vaccine. Data were collected two weeks apart following each dose of vaccination. Collected data were entered in Microsoft Excel 2019 MSO (Version 2021). The presence or absence of symptoms was indicated as 1 or 0. If any symptom was observed within the total duration of the study, we marked it as 1 for symptom seen, and 0 for not seen. Any specific post-vaccination effects seen over any duration were marked as observed. Data were then analyzed statistically using Statistical Package for Social Sciences (SPSS Version 2022). The normality of data was tested using the Shapiro-Wilk test. The Chi-Square test was used to observe the association between the categorical variables and symptoms of 1st and 2nd doses of the vaccination. The associations between the categorical variables and individual symptoms were also studied. Also, to comprehend the association between the frequency of vaccination (1^st^ and 2^nd^ dose) and symptoms presented within the duration of the study, the Wilcoxon Signed Rank test was implemented. The evaluated data were presented in the form of tables and the association is considered to be significant if the P-value is less than 0.05.

## RESULTS

A total of 156 participants were included in the study, the majority of participants consisted of males (66.7%), primarily comprising intern doctors within the age group of 21-24 years. (**Table 1**)

**Table 1.** Demographic parameters.

| S.N. | Parameter | No. (%) |
| --- | --- | --- |
| 1. | **Gender** | Male 104 (66.7%)<br>Female 52 (33.3%) |
| 2. | **Year of Study** | Third year 38 (24.4%)<br>Final year 57 (36.5%)<br>Intern doctors 61 (39.1%) |
| 3. | **Age** | 21-24 yrs. of age 98 (62.8%)<br>25-28 yrs. of age 58 (37.2%) |

About 80.1% of the participants exhibited one or more symptoms after the first dose, which decreased to 66% after the second dose of vaccination. Both doses of the vaccination demonstrated a greater effect in males with the effect being more pronounced following the 2^nd^ dose. Females also exhibited a response, although not as significant as that observed in males. (**Table 2**)

**Table 2.** Symptoms following the first and second dose of vaccination

| Dose | Symptoms | Male (%) | Female (%) | Total (%) |
| --- | --- | --- | --- | --- |
| 1<sup>st</sup> Dose | Seen | 87 (69.6) | 38(30.4) | 125 (80.1) |
|  | Not Seen | 17(57.8) | 14 (45.2) | 31 (19.9) |
| 2<sup>nd</sup> Dose | Seen | 76 (73.8) | 27 (26.2) | 103 (66) |
|  | Not Seen | 28 (52.8) | 25(47.2) | 53 (34) |

The predominant symptoms were pain at the injection site, myalgia, fatigue, itching, and fever. Pain at the injection site was the most commonly reported side effect (48.1 %) accompanied by myalgia (29.5 %) following 1^st^ dose of the vaccination. However, in 2^nd^ dose, pain at the injection site (48.7%) was subsequently followed by fatigue (16%). The emergence of the sore throat and runny nose occurred only during the administration of the 2^nd^ dose of the vaccination. Both doses of vaccination yielded unreported side effects which included diarrhea, difficulty in breathing, tingling sensation, or anaphylactic reaction. The vast majority of the symptoms appeared within 12 hours post-vaccination. (**Table 3**)

**Table 3.** Frequency and Duration of Symptoms.

| Symptoms | Dose 1 | Maximum Duration of Appearance | Dose 2 | Maximum Duration of Appearance |
| --- | --- | --- | --- | --- |
| Pain at the Injection site | 75 | 66(Within 12 hour) | 76 | 72(Within 12 hour) |
| Myalgia | 46 | 42(Within 12 hour) | 21 | 17(Within 12 hour) |
| Fatigue | 42 | 34(Within 12 hour) | 25 | 23(Within 12 hour) |
| Itching | 30 | 26(within 12 hour) | 12 | 12(Within 12 hour) |
| Fever | 27 | 19(Within 12 hour) | 11 | 10(Within 12 hour) |
| Headache | 10 | 9(Within 12 hour) | 9 | 8(Within 12 hour) |
| Dizziness | 9 | 8(Within 12 hour) | 1 | 1(Within 12 hour) |
| Skin Rash | 7 | 6(Within 12 hour) | 3 | 3(Within 12 hour) |
| Redness | 4 | 4(Within 12 hour) | Not seen | Not seen |
| Joint Pain | 3 | 3(Within 12 hour) | 4 | 4(Within 12 hour) |
| Nausea and Vomiting | 3 | 3(Within 12-24 hour) | Not seen | Not seen |
| Swelling | 1 | 1(Within 12 hour) | 1 | 1(Within 12 hour) |
| Cough | 1 | 1(Within 24-48 hour) | 3 | 3(Within 12 hour) |
| Sneezing | 1 | 1(Within 24-48 hour) | 1 | 1(Within 12-24 hour) |
| Sore Throat | Not seen | Not seen | 4 | 4(Within 12 hour) |
| Runny Nose | Not seen | Not seen | 1 | 1(Within 12-24 hour) |
| Diarrhea | Not seen | Not seen | Not seen | Not seen |
| Difficulty in breathing | Not seen | Not seen | Not seen | Not seen |
| Tingling Sensation | Not seen | Not seen | Not seen | Not seen |
| Swollen Lymph Node | Not seen | Not seen | Not seen | Not seen |
| Fainting | Not seen | Not seen | Not seen | Not seen |
| Anaphylaxis | Not seen | Not seen | Not seen | Not seen |

The frequently encountered post-vaccination symptoms were more prevalent in males than in females, and the occurrence of post-vaccination pain at the injection site was more frequently observed in males following the 2^nd^ dose. With the exception of pain at the injection site, all other major symptoms exhibited a declining trend subsequent to the vaccination. (**Supplementary file, Table 1**)

The normality of data distribution was tested using the Shapiro-Wilk test. Our data were non-normal in distribution. Chi-square test was used to see the association between the overall symptoms observed post-vaccination with determinant variables (age, sex and years of study). Among all the tested variables, the post-vaccination side effects after 2^nd^ dose showed a significant association with sex, in contrast to the 1^st^ dose. (**Table 4**) We conducted an analysis of the major symptoms observed after both doses of vaccination. Our research revealed a significant link between sex and fever after the first dose. Furthermore, we found a significant difference in pain at the injection site and myalgia following the first dose among two age groups (<25 years and >25 years). **(Supplementary file, Table 2 and Table 3)**

**Table 4.** Variables associated with overall symptoms post-vaccination.

| Variables | 1st Dose | 2nd Dose |
| --- | --- | --- |
| Age | 0.205 | 0.389 |
| Sex | 0.09 | <0.05 |
| Years of Study | 0.52 | 0.09 |

Further, the association was delineated using Wilcoxon Signed Rank test. Major Symptoms that occurred were analyzed for the association with subsequent doses of vaccination. Interestingly there was no such association between pain at the injection site with the doses of vaccination (p-value >0.05) though it was the major symptom recorded in our study. Other symptoms like myalgia, fatigue, itching, sore throat, and fever were found to be significantly associated with the subsequent doses of vaccination with p-value less than 0.05. (**Table 5**)

**Table 5.** Association of major Symptoms with subsequent doses of vaccination.

| Symptoms | P-value |
| --- | --- |
| Pain at the injection site | 0.7 |
| Myalgia | <0.05 |
| Fatigue | <0.05 |
| Itching | <0.05 |
| Fever | <0.05 |
| Headache | 0.8 |
| Sore Throat | <0.05 |
| Runny nose | 0.3 |

## Discussion

Subsequent to the vaccination received, the majority of the symptoms were reported among the age group 21-24 years, with a comparable observation in the study conducted following Covishield vaccination in Nepal where the study reported maximum symptom presentation among individuals aged 18-30 years. (22) The greatest number of adverse effects was predominantly reported among the male participants post-vaccination with 69.6% in the first (1^st^) dose and 73.8% in the second (2^nd^) dose. Contrary to our study, the findings were more pronounced in the females in the study conducted in Sinopharm (Vero cell) at Patan, Nepal, and in the United Arab Emirates. (23,24) The same study was conducted examining the adverse effect of the ChAdOx1 nCoV-19 (Covishield) vaccine which again revealed a higher preponderance in females. (22,25)

In a study involving 156 participants, 80.1% and 66% of the total participants presented with at least one symptom subsequent to the first(1^st^) and second(2^nd^) dose of vaccination respectively whereas the observed effects are more pronounced in other vaccines including the Pfizer-BioNTech vaccine conducted in Jordan having 83.7% in the1^st^ dose and 88.7% in the 2^nd^ dose and ChAdOx1 nCoV-19 (Covishield) vaccine conducted in Nepal reported 91.6%, as opposed to ours. Similarly, Moderna and Pfizer also demonstrated a greater effect after the second dose than the first. (25–27) However, the effect of the Vero cell vaccination in a study conducted at Patan, demonstrates a lesser effect exhibiting 19% in 1^st^ dose and 11.2% in 2^nd^ dose. (24) A notable decrease in the observed effect is seen from 80.1% in the 1^st^ dose to 66% in the 2^nd^ dose in our study. A similar pattern of effect was seen with Vero cell and Covishield in the study conducted in Nepal. (22,24) In contrast to our study, a reciprocal relationship was observed between the dose and the symptoms in a study conducted in Sinopharm (Vero Cell) by Balsam et.al.(23) Similarly, adverse effect was also observed to be increased from the 1^st^ dose to the 2^nd^ dose with Pfizer-BioNTech, AstraZeneca, and Sinopharm (Vero cell) vaccines in the study conducted in Jordan. (26) The most pronounced symptoms were observed within 12 hours following vaccination which is in coherence with the study conducted by Poudel et.al.(22)

Among the 156 participants included in the study, Pain at the injection site emerged as the predominant symptom constituting about 60% of the reported presentation following 1^st^ dose of the vaccination and 73.7% following the 2^nd^ dose. The outcome of our study was consistent with the findings conducted among the HCWs of Jordan which illustrates that pain at the injection site was the most common presentation following both doses of vaccination. (26) A similarly significant finding with Vero cell was seen in a study conducted in Patan, Nepal, and in the United Arab Emirates where again pain at the injection site was found to be the most common presenting symptom. (23,24) Based on the result of the study by the World Health Organization, Pain at the injection site once again emerged as the prevailing symptom (≥1 in 10). (28)Muscle pain or myalgia ranked as the second most prevalent symptom with an average of 58.71% following both doses. Conversely, it contradicts from the study conducted by Balsam et.al where only 6.3% at the 1^st^ dose and 5.9% at the 2^nd^ dose of the total study population presented with myalgia. (23) On average, 58.71% % of the participants presented with fatigue which is regarded as the 3^rd^ most common symptom presentation in our study which diverges from the study conducted by Osama Abu-Hammad et.al and Balsam et.al.(23,26) The manifestation of headache was more pronounced in our study, accounting for 8% in the 1^st^ dose and 8.7% in the 2^nd^ dose, whereas it is less prevalent in the study conducted at Patan, Nepal accounting for 2.9% in the 1st dose and 1.6% in the 2^nd^ dose. (24) However, in the study on Vero cell conducted by Balsam et.al, the headache was found to be 9.6% in the 1^st^ dose and 10% in the 2^nd^ dose. (23) On average, 33.33% % of the total participants exhibited fever which exceeds the average number of presentations in the study conducted by Osama Abu-Hammad et.al and Balsam et.al. (23,26)

In a developing country like Nepal with finite resources, the Vero cell has proven to be the most suitable one because of its easy accessibility, availability, and good efficacy. (29) Correlating our study with other studies, Pain at the injection site, Myalgia and Fatigue were found to be the top three commonest presentations whilst Pain at the injection site was commonest among all the studies. The least common symptoms included swelling, sneezing, sore throat, nausea, vomiting, dizziness, cough, and runny nose. (24,30–32)

In a study conducted at Patan, Nepal, we discovered a significant association between sex and symptoms after both doses of vaccination, but not with age. This pattern was repeated following the Covishield vaccination, which again showed an association between sex and symptoms. Despite the differences between the vaccine used, the results were consistent. Our study indicated that the symptoms were more strongly associated with sex following the 2^nd^ dose than the 1^st^, but not with age and years of study. In a study conducted at Jordan, Pfizer, Sinopharm, and AstraZeneca combinedly showed a significant association with age only after the 1^st^ dose whereas there was no significant association with sex. (24–26)

Upon analyzing the individual symptoms, in our study we found that gender and fever were significantly associated after the 1^st^ dose, but no at any other symptoms. Pain at the injection site and fever was found to be significantly associated with age following the 1^st^ dose of the vaccination. But no significant differences were observed following the 2^nd^ dose. In contrast to our findings, a study conducted by Balsam et.al showed that gender was significantly associated with fatigue after the 1^st^ dose, while the 2^nd^ dose was found to be associated with pain at the injection site and fatigue. Similarly, age showed a significant association with pain at the injection site, nausea, and myalgia following the 1^st^ dose, whereas only fatigue was found to be significantly associated following the 2^nd^ dose. Contradictory, to the overall findings, age and sex were not significantly associated with individual symptoms in a study conducted by Osama Abu-Hammad et.al.(23,26)

## Conclusion

During the peak of the COVID-19 outbreak, when its impact was most severe and survival was paramount, the emergence of vaccine proved to be a lifesaving breakthrough, offering a critical ray of hope. The vaccine has always been a boon to human existence and a pivotal turning point in the survival of mankind. The effectiveness of the vaccine requires no further validation.

Based on our comprehensive study, following both doses of vaccination, Pain at the injection was found to be the commonest presentation which carried no long-term side effects, and all the symptoms resolved within 72 hours of presentation.

This is precisely why Sinopharm (Vero Cell) is highly favored, as it demonstrates remarkable efficacy without compromising on affordability in a developing country like NEPAL. Nevertheless, a larger study on the general population including all age groups is highly recommended to detect all of the side effects.

## Data Availability

All data produced in the present study are available upon reasonable request to the authors

## AUTHOR CONTRIBUTIONS

**Rupak KC:** Conceptualization; ethical approval; data collection; software; writing-original draft preparation; supervision; writing-review & editing. **Sibika Malla:** Resources; methodology; data collection; writing-original draft preparation; writing-review & editing. **Aashish Pandey:** Methodology, software, writing-original draft preparation; writing-review & editing. **Merina Shrestha:** Investigation; resources; ethical approval; supervision. **Saharoj Siddiqui:** writing-original draft preparation; writing-review & editing **Suraj Adhikari:** Project administration; resources; software; writing-original draft preparation; supervision; writing-review & editing. **Niranjan KC:** Project administration; data collection; resources; writing-review & editing. **Rumi KC:** Resources; data collection; project administration; writing-review & editing.

## ACKNOWLEDGEMENT

We would like to acknowledge Dr. Prabesh Pant, Assistant Professor, Department of Neuro-medicine, COMS, Chitwan, Banke for his invaluable support and guidance. We would like to acknowledge Dr. Dhan Bahadur Shrestha for his cooperation and valuable insight into this paper. We would also like to express our sincerest gratitude towards our participants for their time and co-operation.

## LIMITATIONS

Our study was conducted in a small center where the overall population was not included. All the spectra of side effects of the vaccine in a healthy group population were difficult to appreciate. Also, recall bias was significant.

## CONFLICT OF INTEREST STATEMENT

No any supporting source or financial relationship has been involved throughout the study. The authors declare no conflict of interest.

## DATA AVAILABILITY STATEMENT

The authors confirm that the data supporting the findings of this study are available within the article and its supplementary materials.

## ETHICS STATEMENT

Ethics approval for the research project was obtained from the NGMCTH Ethics Review Committee (Ref.463/078-079) on 29 December 2021.

## TRANSPARENCY STATEMENT

The lead author Rupak KC affirms that this manuscript is an honest, accurate, and transparent account of the study being reported; that no important aspects of the study have been omitted; and that any discrepancies from the study as planned (and, if relevant, registered) have been explained.

All co-authors have read and approved the final version of the manuscript. The lead author Rupak KC had full access to all of the data in this study and takes complete responsibility for the integrity of the data and the accuracy of the data analysis.

## Notes

### Competing Interest Statement

The authors have declared no competing interest.

### Funding Statement

This study did not receive any funding

### Author Declarations

IRC of Nepalgunj Medical College gave ethical approval for this work.

### Summary of Updates

We revisited and re-entered the data after being rejected at PLOS One journal due to simple statistics. We have now analyzed the data and made associations with categorical variables with symptoms, also symptoms with the frequency of vaccination were also analyzed.

