## Supplemental Table 1 for "“Side effects of Vero cell vaccination against Covid-19 among medical students of Nepalgunj Medical College”-A Post Vaccination survey"

**Table 1. The prevalence of post-vaccination symptoms seen in both gender.**

| Symptom | Dose | Male | Female |
| --- | --- | --- | --- |
| Pain at the injection site | 1st dose | 53 | 22 |
|  | 2nd dose | 55 | 21 |
| Myalgia | 1st dose | 31 | 15 |
|  | 2nd dose | 16 | 5 |
| Fatigue | 1st dose | 32 | 10 |
|  | 2nd dose | 18 | 7 |
| Itching | 1st dose | 25 | 5 |
|  | 2nd dose | 10 | 2 |

**Table 2. Association of individual symptoms with gender.**

| Symptoms | 1st Dose | 2nd Dose |
| --- | --- | --- |
| Pain at the injection site | 0.19 | 0.09 |
| Myalgia | 0.53 | 0.23 |
| Fever | **0.05** | 0.07 |
| Fatigue | 0.89 | 0.36 |
| Itching | 0.23 | 0.17 |
| Headache | 0.21 | 0.37 |

**Table 3. Association of individual Symptoms with age interval (less than 25 and more than 25).**

| Symptoms | 1st Dose | 2nd Dose |
| --- | --- | --- |
| Pain at the injection site | **0.007** | 0.532 |
| Myalgia | 0.191 | 0.97 |
| Fever | **0.003** | 0.36 |
| Fatigue | 0.368 | 0.542 |
| Itching | 0.283 | 0.31 |
| Headache | 0.6 | 0.204 |
